# Migrant healthcare workers’ experiences and mobility patterns in a global health crisis: a qualitative study with Romania physicians working in Germany

**DOI:** 10.1101/2023.04.29.23289300

**Authors:** Ellen Kuhlmann, Marius-Ionuț Ungureanu, Nancy Thilo, Leonie Mac Fehr, Nicoleta-Carmen Cosma, Monica Georgina Brînzac, Alexandra Dopfer-Jablonka

**Author notes:** Corresponding author, Dr Ellen Kuhlmann, Department of Rheumatology and Immunology, Hannover Medical School, Carl-Neuberg-Straße 1, 30625 Hannover, Germany. Ellen Kuhlmann and Marius-Ionut Ungureanu share first authorship.

## Abstract

**Introduction:** Attention to the healthcare workforce has increased, yet comprehensive information on migrant healthcare workers is missing. This study focuses on migrant healthcare workers’ experiences and explores mobility patterns in the middle of a global health crisis, aiming to explore the capacity for circular migration and support effective equitable healthcare workforce policy.

**Methods:** Romanian physicians in Germany served as an empirical case study. We applied a qualitative explorative approach; interviews (n=21) were collected from mid of September to early November 2022 and content analysis was performed.

**Results:** Migrant physicians showed strong resilience during the COVID-19 crisis and rarely complained. Commitment to high professional standards and career development were major pull factors towards Germany, while perceptions of limited career choices, nepotism and corruption in Romania caused strong push mechanisms. We identified two major mobility patterns that may support circular migration policies: well-integrated physicians with a wish to give something back to their home country, and mobile cosmopolitan physicians who flexibly balance career opportunities and personal/family interests.

**Conclusions:** Health policy must establish systematic monitoring of the migrant HCWF including actor-centred approaches, support integration in destination countries as well as health system development in sending countries, and invest in evidence-based circular migration policy.

## Introduction

WHO has declared the healthcare workforce (HCWF) a global policy priority (1, 2) and efforts have been stepped up in the European region to highlight the threats of growing health labour market shortages and increasing workload, stress and mental health risks of healthcare workers (HCWs), exposed to increasing violence (3, 4, 5, 6, 7, 8, 9, 10). However, data on the migrant HCWF are still poor (11, 12, 13, 14, 15, 16), especially concerning the particularities of intra-European Union (EU) mobility (17, 18), and information on needs and perceptions of individual HCWs is largely lacking (19). We do not know how migrant HCWs experienced the COVID-19 crisis and how it might shape mobility decisions.

The lack of data on migrant HCWs reveals a serious gap in HCWF policy and research. Migrant HCWs account for a relevant share of the HCWF (20) especially in high-income countries. Growing health labour market shortages and post-pandemic retention challenges in an already stressed and burned-out HCWF (3, 21) together with economic hardship in sending countries are likely to further accelerate migration flows (22). WHO reports an increase in the international recruitment of HCWS and highlights the negative impact on health systems that are losing their HCWs (23). In Europe, international recruitment was identified as an increasing source of new HCWs, that may in some countries and professional groups even exceed the number of domestic graduates, as observed in medicine (3).

An increase in migration happened in a time of extraordinary hardship for both the individual HCWs and the healthcare systems, calling for urgent action to develop more equitable HCWF governance beyond the voluntary tools of the global Code (24, 25, 26). Against this backdrop, ‘circular migration’ may gain new currency, although the concept is neither new nor without criticism (27, 28, 29, 30, 31, 32, 33, 34). It comprises all forms of mobility flows between two or more countries that exchange larger groups of HCWs, aiming to improve flexibility for HCWs and legal support for mobility back and forth between countries (13, 14, 35, 36, 37).

Circular migration is supposed to create a ‘triple win’ – for the sending country, the destination country and the individual HCWs, yet an EU study concluded in 2016 that the approach was rarely used (36, 37). Until today, it seems to be the least field-tested tool of HCWF policy (38). However, receiving countries with high-resourced healthcare systems seem to be more positive about cross-country, usually bilateral mobility agreements than less well-resourced and sending countries, as can be observed in Germany (39). A lack of comprehensive monitoring may nurture interest-driven policy and exacerbate European and global inequalities thus calling for urgent action due to HCWF shortages and the COVID-19 challenges.

Our study seeks to contribute new knowledge in support of equitable and effective solutions to HCWF policy and governance, with a focus on Europe. We introduce an actor-centred approach to explore perceptions in the middle of a global health crisis and emergent mobility patterns that embody the capacity for ‘circular migration’ from the bottom-up. Germany and the group of Romanian physicians were chosen for an empirical investigation, drawing on qualitative interview methodology.

## Methods

### Research design and country case

We applied an explorative qualitative approach based on semi-structured interviews and informed by an intersectional research framework that places individual migrant HCWs in the context of health systems, organisations, and professions (40). We set the focus on EU mobility and actor-centred perceptions of the COVID-19 situation. Romanian physicians in Germany make for an interesting case study that may illustrate typical conditions and problems of the Central and Eastern Europe (CEE) countries and EU health labour market policy (35, 41). Notwithstanding a ‘huge dependence on health workers from outside the European region’ (42) creating a gap in health systems in Europe (14, 26, 29), the intra-EU mobility embodies specific problems, as well as opportunities for novel health workforce governance approaches (41).

#### Box 1.

The case study: health workforce figures in Germany and Romania

Germany is an example of a high-income country that managed the pandemic well (43) and has an overall high HCWF density with a ratio of 4.53 physicians to 1,000 populations (20). At the same time, the pandemic has worsened the shortage of HCWs and work conditions [6]. High-income countries show a generally higher share of migrant HCWs that is likely to increase in the future (3, 44). The number of annual medical graduates in Germany is below the EU average (3). Foreign-trained HCWs account for 12.0% and nurses for 7.1% (20); health policy is currently scaling-up efforts to recruit migrant HCWs (39, 45). Physicians born/trained in Romania are supposed to be the largest group of migrant physicians in Germany; numbers increased strongly over recent years (46).

Romania is an example of the CEE countries that strongly suffered from COVID-19 and did so ‘with far fewer health workers than they needed’ (5); these conditions increased stress and burn-out syndromes of the HCWs in the (47). ‘Many of those who could have helped had already moved to the West’ (5), thus unmasking the EU’s health labour market and policy failure during the pandemic. Romania scores below the EU average of HCWF density (3), with a ratio of 3.33 physicians to 1,000 populations (48). The country is struggling with high outflows for years (25, 49); anecdotal evidence suggests a huge outflow to Germany. However, the number of annual medical graduates is far above the EU average (3), which may motivate migration and further increase the outflows.

According to WHO Europe, no reliable data are available on the outflows of HCWs (3) and coherent categories and reporting systems are missing (13). Thus, some clarification of the terms may be useful. Existing public statistics are mainly based on two categories: the number of medical and nursing graduates disaggregated for foreign-trained and domestic-trained, and the number of requests for certificates of qualifications as required by destination countries (3, 20). Occasionally, data on foreign-born and domestic- (or national-) trained HCWs may be available from surveys. In our study, we use ‘migrant’ HCWs as an umbrella category for foreign-born and foreign-trained HCWs and ‘mobility’ and ‘’migration’ as interchangeable terms, regardless of the country of origin. In legal terms, employment moves within the European Union are a specific case based on free movement law as a pillar of EU law (18).

### Data collection and analysis

Following ethical approval, we invited physicians to participate in a qualitative interview using two different channels: snowball sampling initiated via existing contacts in Germany and Romania, and random selection via publicly available register data and information on ambulatory care physicians. The majority of physicians in our sample were recruited via snowballing. Next to a very low response rate, the random selection created a strong bias against older physicians with longstanding work experience and German citizenship.

Our sample sought to reflect the complexity of the migrant category and the specific situation of the CEE countries and Germany. Most interviewees were born in Romania and moved to Germany as EU citizens, but a few other cases were also considered: trained but not born in Romania; born in Moldova and trained in Moldova and Romania; trained in Romania during the time of the communist regime and moved back to the former GDR after the fall; born in Romania and moved to Germany as child/young adult during the time of the communist regime. Table 1 provides an overview of the composition of the sample (n=21), ensuring sufficiently anonymised information.

**Table 1.** The composition of the sample

| Item | Participants |
| --- | --- |
| Region | Geographical diversity including East and West regions, but some clustering in North Rhine Westfalia, Lower Saxony and Baden-Württemberg |
| Gender | 13 women, 8 men (as identified by participants) |
| Workplace information | 5 university medical centres.<br>12 hospitals (without universities) including smaller/larger and public/private provider organisations (large chains, churches, and small private owners).<br>4 ambulatory care, single practice, shared practice owners, employees. |
| Professional status | Academic status ranged from high-level international expert, middle-level and junior physician; occupational position considered several residents (some in their final stage), junior physicians, senior physicians with leadership, 1 women physician on maternity leave and 1 pregnant with employment ban due to COVID-19 infection risk. |
| Profession, speciality | General medicine, internal medicine (several specialised areas), anaesthesiology, surgery (several specialised areas), and three smaller specialities including ophthalmology, psychiatry and radiology. |
| Personal situation | All, except one, physicians were living in partnership/married including with partners from Romania, Germany, or other EU countries; most had children; two had commuter models with their partners in Romania. |

Source: authors' own table

The interviews were carried out in German between mid of September and early November 2022, using a semi-structured topic guide developed by the German-Romanian project team. The topic guide comprised four key issues: work and employment situation in Germany; experiences during the COVID-9 pandemic; reasons for migrating to Germany, and future plans; it was flexibly applied and adapted to the interview situation. Most interviews were carried out via video call, one via phone and another face-to-face, lasting on average one hour, from 20 minutes to nearly two hours. Most were audio-recorded and subsequently transcribed (notes were taken in a few cases of technical problems) and additionally, reflective protocols were prepared.

The data analysis was informed by qualitative content analysis. An on-site workshop was organised mid of October 2022 in Cluj-Napoca, Romania, to discuss the framework and preliminary findings; additionally, feedback and comments were gathered during a public seminar. The analytical framework proofed to be sufficiently robust and was applied to the entire interview sample. Data saturation was achieved with 21 interviews included in the analysis, assisted by the MAXQDA (version 2020) qualitative data software. Selected quotes from the interview were translated into English.

## Results

### Work and employment situation

Our interview partners mainly highlighted positive work and employment experiences and some were enthusiastic about their situation. “To be honest, I am extremely satisfied right now” (#10, male [M]). “The atmosphere was always perfect and very nice” (#19, female [F]), and “it is great here in this hospital” (#4, F), were some of the descriptions. The individual stories certainly varied greatly and there were also more critical issues, yet we observed in almost all interviews an underlying pattern of foregrounding positive experiences and downplaying negative ones. This ‘pink lens’ showed up in different ways. One important strategy was linking negative experiences to the situation as a newcomer in the country, sometimes reinforced through a lack of experience as a physician.

> Yes, and originally, I didn’t know any German, so it was a very jerky beginning, I thought. And I don’t have such fond memories of the time in surgery. I was pretty much thrown into cold water there. (#9, F)

Another mechanism was portraying problems as part of the job that affect everybody in the same way, but can be handled through strong motivation and/or team support.

> Sometimes you don’t have a break – then you’re totally exhausted. I don’t agree with that either, but that’s a totally different issue. So, this work overload, this understaffing always. I have the impression that we are always understaffed, but if it’s fun and if you do it with a clear conscience, then it’s okay. (#8, F)

A third important way of coping with problems was the comparison with other hospitals and workplaces, or with the situation in Romania. “That’s why I’ve always said that I think it’s great here in this hospital, I like it very much, because I’ve also worked elsewhere.” (#4, F)

Viewing the own situation through a pink lens did not mean ignoring the existing HCWF problems and may even co-exist even with harsh criticism of the systemic conditions.

> “That’s not possible, you have to put the brakes on this capitalism somehow and you have to employ enough staff and not always say ‘operate more and more’, ‘more complicated operations’, ‘increase the revenue’.” (#5, M)

The physicians in our study addressed commonly known and widely criticized HCWF problems (e.g. Buchan et al., 2021; Kuhlmann et al., 2021; WHO Euro, 2022 time to act). High workload, long work hours, no regular breaks, and too little time for family and leisure activities, as well as to little time for patients, were major issues. Younger physicians furthermore highlighted a lack of mentoring and poorly organised resident training, “so, that it needs a lot of individual initiative” (#10, M), and mainly women mentioned the problems of combining family life/childcare and medical practice and/or an academic career. “Well, I’ve always loved it and I would like to stay in the clinic. …But it has to remain family-friendly, at least as it is now…so that I can work in the clinic with four children” (#2, F). They tried to negotiate more flexible work conditions and some changed their employer.

### The needs of migrant physicians

We found only a few hints of needs specifically related to the situation as a foreigner, except for the language barriers. If needs were mentioned, they were mainly related to the transition period to Germany.

> I wish I had had that [support] before. Now I don’t need it any more. The beginning was made really difficult for me here [in Germany]. I would have needed more support, even without Corona… Now [current surgery] it is very good. … It’s really very good there. (F, #1)

Other important issues were problems stemming from differences in the education and training systems. In Romania, physicians get their approbation immediately after their exams, while a ‘practical year (PJ)’ is mandatory in Germany. “The [PJs, students in their Practical Year] learn all the things. And in Romania there is no such year. It’s a bit difficult to explain” (#6, F). Another physician reported that he and his wife wanted to participate in the PJ training but were denied access; he felt that German PJ students got more attention from the lead doctors than the young Romanian doctors, which he perceived as discrimination.

On the positive side, the benefits of role models and mentoring programmes were highlighted.

> That helps me a lot as a migrant, because I … there is so much information that you don’t know at all in such a structure. And such a programme [mentoring] can help us as migrants, but also women as a whole, to get a bit of an overview. (#15, F)

### The COVID-19 pandemic situation

The workload and stress were very high: “We often don’t have a real break. So, sometimes we work twelve hours and even at the end up to 24 hours” (#18, M), yet protection was perceived overall positive: “So, we got plenty of masks, protective clothing. And we certainly got gloves and all kinds of things. It was not a bottleneck at all, at least not for us” (#11, M). Shortage of personal protective equipment (PPE), where it happened, was a problem in the early days of the pandemic and “affected everybody because nobody was prepared … Staff had to bring their own masks“ (F, #1). The COVID-19 vaccine was generally perceived as very helpful, yet one physician felt to be put under pressure by the compulsory vaccination.

We observed a common narrative of high workload but collegial support during the pandemic and did not find disadvantages related to the status as a foreigner. Several interviewees highlighted that the COVID-19 situation affected everybody in the same way and that it has improved after the first wave.

> Yes, colleagues were ill and then we had to cover for them a bit more each time. And that was very tense. Yes. But yes. Somehow we solved the situation. Whether I had support myself or asked for support? I don’t remember. Yes, sure. We supported each other, but I don’t have any, no special, bad or good experiences there. (#15, F)

Many participants in our study had very strong connections with family and friends in Romania that were disrupted and limited to virtual social media during the pandemic. Here, too, we observed a strong attitude of not complaining and a high commitment to prevention and protection rules.

### Social relations and discriminatory experiences

A positive attitude served as a kind of master file that shaped the perceptions of work experiences, the relationships with colleagues and copying with discriminatory experiences. Several interviewees highlighted they did not experience disadvantages at work. “I was lucky, I had colleagues who immediately wanted to show me how to do this and that, and they always helped me” (#19, F). “I never felt disadvantaged or so, moreover I was lucky and got a leadership position very early” (#3, M). “Not at work, at work I have little experience [with discrimination]” (#8, F). Discrimination experiences, if mentioned, were usually related to a transition period as a newcomer or excused with a high workload of colleagues.

> “And I’m not sure, I always have to ask. And sometimes my colleagues are also busy and they don’t have… so I can understand, they don’t have so much time and so much patience to explain everything and that’s not so good for me, so to speak…Sometimes it’s all right. But sometimes there are so… it’s sometimes difficult to communicate with some people” (#6, F)
>
> “Discriminated, if I use the harsh word, the harsh word, but I think you know what I mean. So, I never felt any disadvantage because I am a foreigner. Really, never, never, except for maybe at the very, very beginning. I started in Thuringia. I met some nice people there, but … I was a newcomer to the profession and unsure of myself. (#5, M)

Discrimination was more clearly addressed if it was related to patients and their relatives rather than to colleagues, as well as to the private sphere, in particular, to previous employment in the Eastern part of Germany. One participant mentioned that her professional status as a physician was not respected by lower-status professionals and insisted on the professional hierarchy. She did not experience discrimination from colleagues, but described nurses “as our enemies” (#4, F) and felt inappropriate task distribution by secretarial staff. Different attitudes on professional hierarchies and interprofessional practice, as well as gender-based role conflicts, might create perceptions of discrimination.

A women physician explicitly addressed the intersectionality of discriminatory experiences, highlighting persisting gender-based discrimination that may be stronger and more violating than the status as a foreigner.

### The push and pull factors of migration and future plans

The reasons for leaving Romania and choosing Germany as their destination country were shaped by complex individual conditions and the political situation at the different times of migration decisions that we considered in our sample. However, we identified some common patterns of push and pull factors. On the push side, four major drivers emerged.

- A poorly resourced healthcare system
- Poor career opportunities and lack of choice.
- Nepotism and lack of transparency and fairness in the promotion process.
- Widespread corruption in all areas of the healthcare system, academia, and society.

These reasons were often interconnected and motivated a decision to leave the country. As one physician put it: “We have a lack of everything [in Romania], and then there is the corruption” (#5, M). Professional and health system-related aspects appeared to be strong drivers for migration. Personal aspects – having a better life, higher salaries, family members in Germany, or a better future for children – were sometimes mentioned but generally seemed to play a weaker role. A younger physician highlighted that money was not an issue, because their financial situation in Romania was not bad and salaries had generally improved over recent years.

We found a corresponding pattern of push factors.

- High-tech medicine and high medical standards provide better opportunities for improving medical knowledge and technical skills.
- Opportunities for professional and career development, especially for young physicians.
- Residency training and the freedom of choice of the medical speciality.

Professional standards and institutional conditions of the health and education systems were major reasons to choose Germany, sometimes coupled with better living conditions and higher salaries. It should be mentioned that we did not find any clear sign of gender-based differences in motivational factors for migration. Table 2 provides some examples; one participant brought it to the point:

**Table 2.** Push and pull factors motivating migration decisions

| Romania, push factors | Germany, pull factors |
| --- | --- |
| And in Romania we don't have that many options. So there are the big hospitals in the big cities ... but there are by far too many people, and also the corruption, that...and also after the residency, we can't find a job as a specialist. ... There is the son of a chief physician and he gets the job, although I am better qualified, and then I am waiting five years and there is another relative who is outflanking me." (#6, F) | Professionally, where I am, it's such a high-level medicine, and I've always liked that, always loved that. And that was actually always the reason why I came here. (#2, F) |
| I thought that I would like to offer the best possible therapy, and in Romania, this is often not possible because of costs that are not covered or different modern therapies that are not offered at all. (#19, F) | And here you can see a case from A to Z in the hospital, so in the sense the patient comes and you do all the examinations and he leaves with a diagnosis, and everything is clear in Germany. In Romania, the patient still has to get things himself, in two or three months he has a CT scan and then we wait. You can't really learn because you don't get to see it. (#4, F) |
| But that is difficult in Romania. You can't find a place [as a resident] like you can in Germany, you're pretty much tied to the starting position [which is assigned according to exam grade], at least as an assistant doctor. (#21, M) | That it was completely different in Romania. ... the salaries were much lower and our examination system doesn't allow you to choose your desired speciality, but you are categorised like this, depending on your grade. (#9, F) |
| These chief physicians are still from the communist era, they are still old, they are still corrupt. And the money they get for equipping the hospital, they use for their own practices. (#4, F) |  |

> And then, Germany gave me the alternative, yes: Do you absolutely want to learn medicine, here you go. If you want to work by all means, here you go. If you want to earn, you can earn. If you want to earn more, then you have to work more, and, um, yes. (#8, F)

### Future plans and emergent mobility patterns

The future plans were more diverse and flexible than the factors that motivated migration and the choice of the destination country. Personal, professional, and system-related reasons were also more strongly interconnected. Next to the workplace and work conditions, as an employee or self-employed surgery owner, the living conditions, family relations, socio-cultural context and political conditions, especially the role of right-wing populist parties and anti-migrant policies, were important criteria for staying or leaving, while money was not a key issue. We identified four major types of mobility:

- the mobile cosmopolitan physician,
- the integrated physician with an open future,
- the settled-in-Germany physician,
- the wishing-to-return physician.

### The mobile cosmopolitan physician

This type was usually a younger physician (sometimes with several years of work experience in Germany) employed in a hospital with strong professional motivation and ambitious career goals. These physicians were highly flexible and aimed for the best work and living conditions. For instance, one participant said, she feels like ‘a European citizen’, no longer as Romanian but also not as German (#7, F). She sought to go where she likes it most and did not constrain her career choices by nationality; she also had a strong interest in global health including working abroad outside the EU. Another physician was since longer practising a commuter model to combine advantageous work conditions in Germany with family life in Romania. A third one went to Romania for medical education and to Germany for residency training and was considering to Switzerland after residency or back to his home country Italy. Thus, the conditions were highly diverse but mobility and a cosmopolitan spirit emerged as a common denominator.

### The integrated physician with an open future

In this group, we found older and younger women and men physicians, mainly with children and working in the hospital sector. They had a preference for staying in Germany; in some cases, a clear decision was limited to residency training at the first instance, or dependent on the political climate and future impact of right-wing parties and movements: “I watch what happens. And if I get the impression that my child is in danger or that I am in danger or something… then I would also return home” (#8, F).

Characteristically, this type of physician was ambiguous about their migration decision and had strong emotions to ‘keep a door open’ to Romania and ‘give their country a chance’.

> So that’s always…, I think about it almost every week: How would it be to go back to Romania? Because I felt more useful there than in this society. Because, as I said, I always had this willingness: we have to change something, we have to change a lot in the health system [in Romania]. There are a lot of problems, but here [in Germany] it works reasonably well. (#14, F)
>
> Initially, the plan was actually, I come, I do the residency here in this super big hospital, where you have everything, and then I go back. So that I then also want to bring my support home. It’s a bit like the dream I had back then. Maybe it hasn’t completely disappeared, I have to say, yes. Because at home, in the homeland, good specialists are actually also needed. But now, of course, with the whole family situation, it’s turned out a bit differently, and so. (#2, F)
>
> Mhm, [to go back to Romania is] a need, I would almost say. To give my home country another chance and see if I can also use my professional knowledge here [in Romania]. (#5, M)

### The ‘settled-in-Germany’ physician

High professional commitment, investment in training and language skills, and engagement in the job were key characteristics of this type. Difficulties were perceived as temporary problems and manageable through changing the employer, moving to ambulatory, or partnering with a surgery owner. Some had German citizenship or were considering applying for it. This type comprised a diverse group of physicians. Some were older physicians living in Germany for many years, including in academic leadership positions or as surgery owners, but we also found young physicians in an early stage of their career. The reasons may be related to personal issues: “I’m married, I have a German husband, I have two children, they are also German, so to speak” (F, #9), or “my mum lives in… and I wanted to stay near here” (#6, F), as well as to institutional conditions in Romania.

> That’s a very difficult question because I think most Romanians – and unfortunately I’m one of them – they don’t have confidence in the system there. So I think that if I go back [to Romania], I have no confidence that I will get out of there again. (#19, F)

The decision to stay may also be motivated by a wish ‘to give back’, as described previously for the ‘open future’ type, but the results were different. Some of the surgery owners said they treat many patients from Romania and Eastern European, as well as refugees, and feel responsible to support their Romanian fellows. Others linked the wish to ‘give back‘ to Germany: „If Germany allows me to, I would live here, pay taxes and adapt to the best of my ability to the rules, which are better here than at home” (#8, F).

> So I’ve been in this country much longer now than in the country where I was born. My son grew up here and is really German and loves this country. We have our roots in Romania, I am also very proud of my school, my university, my past. But I would say I am at home here and I am grateful every day for what I have received in this country. And happy to somehow be able to give something back through my deeds. (#16, F)

### The ‘wish-to-go-back’ physician

Only one participant in our study clearly said she wants to go back to Romania, although the work situation and support of colleagues were perceived as very good: “I am ready! Whenever! Even now, I am ready to pack my suitcase. So it depends on my husband“ (#4, F).

## Discussion

Our study set out to put migrant HCWs centre stage and provide deeper insides into perceptions, motivational factors, and mobility patterns. We now apply a policy lens and discuss what lessons can be drawn from our findings for healthcare governance and HCWF policy. It should be mentioned that the migration status intersects with other social categories that shape experiences and eventually inequalities, as the example of gender may illustrate (50, 51, 52), yet these conditions cannot be explained more systematically from our material.

To begin with, the physicians in our study were mostly positive about their mobility decision and their work situation, notably, this includes the COVID-19 period and contemporary experiences. Indeed, these findings came as a surprise in the middle of a most severe global HCWF crisis (6, 7, 53), that has caused several waves of HCW strikes in German hospitals (54) and strong complaints in the ambulatory sector. A closer look at the individual stories revealed some explanations for a pink lens, as we called it.

Downplay difficulties may serve as an effective coping strategy in both the workplace and private sphere, and the crisis situation certainly increased need for coping mechanisms. Furthermore, the physicians showed several attitudes that made them more resilient, not only during the COVID-19 pandemic. They were committed to high medical standards and had a strong professional identity that may improve integration and buffer discrimination experiences (55). They had lower expectations concerning employer and state responsibilities and often compared their situation with Romania where welfare state policy is generally weaker and trust in government low. They had a strong willingness to manage problems and an overall high problem-solving capacity, e.g., through changing the employer, the city, the healthcare sector, or the country. The integration challenges and experiences might have increased their resilience toward stressful situations more generally and boosted a degree of cognitive flexibility.

Resilience and flexibility ensure agency in difficult times; the coping attitudes may be individually effective but bear the risk of silencing migrant HCWs in HCWF policy and hiding governance gaps and policy failure. Migrant HCWs must be included more actively in all areas of decision-making to respond better to individual needs and strengthen health labour market policy. Our examples reveal that poor work and living conditions may create a high turn-over of employers and eventually motivate mobility to other countries, thus impacting negatively retention.

The relationship between health policy and the retention of migrant HCW is far from being clear and needs further investigation. Our findings suggest that the health system and socio-political conditions play a major role in motivating previous and future mobility decisions. Conditions at the organisational level (employer, workplace) seem to have an overall weaker impact, because agency and choice to improve career pathways are generally higher on this level. This mirrors findings from Ireland and the argument of Humphries and colleagues (14) that health system conditions are important factors impacting migration decisions. The organisation of work must therefore be explored within this context (56). Research undertaken with nurses in the English NHS found a higher drop-out rate of EU nurses compared to national and non-EU nurses (57); this may indicate growing competition between EU countries.

We found strong connections between the push and pull factors in sending and destination countries, highlighting a need for transnational HCWF policy efforts. For instance, EU investment in anti-corruption law and the healthcare system in CEE countries may create co-benefits for HCWF retention in sending countries. There were also some more practical and short-term suggestions emerging from our study concerning national macro and meso-level interventions. For instance, in Germany, an opening of the ‘practical year’ programmes for Romanian physicians, improved mentoring, and gender equality programmes and support for women leadership may improve the integration of young migrant physicians. On the other hand, in Romania, innovation in residency training to enable choice of the specialty might reduce outflows and greatly improve recruitment and retention.

Finally, our findings bring novel opportunities for the ‘circular migration’ policy into perspective, that move beyond retention policy based on national interests. The ‘mobile EU citizen’ and the ‘integrated physician with open future’ emerged as the most relevant sources of a flexible transnational exchange of HCWs that have not been explored previously. There are certainly many question marks concerning circular migration, but the need and urgency for action call for new efforts. Here, our findings may provide some guidance on how to build capacity from the bottom-up and develop more participatory and equitable modes of HCWF governance that move beyond interest-driven promises of the circular migration policies promoted by high-income countries.

### Limitations

Our study has several limitations that should be considered. The qualitative material provided in-depth information that allowed us to explore individual perceptions, but no conclusion can be drawn on health labour market trends, COVID-19 risks, and discriminatory practices. Also, relevant groups of migrant physicians from Romanian might be lacking in our sample; we do not know whether the findings apply similarly to physicians from other countries or to other professional groups. More generally, identifying capacity for equitable policy options does not allow conclusions on implementation. The research was designed as a pilot project and may thus provide guidance for future research and systematic monitoring of HCWF migration.

## Conclusions and policy recommendations

We sought to bring the situation of migrant HCWs and an actor-centred approach into the HCWF policy debate and explored the capacity for more equitable HCWF policies. New international efforts to prioritise the HCWF and respond to the crisis are currently creating windows of opportunities for policy change that must still be explored more systematically. Four major conclusions are emerging from our research.

- HCWF policy must pay greater attention to migrant HCWs and actor-centred approaches to close existing knowledge gaps and respond more effectively to both individual and health labour market needs.
- Effective retention policy must consider mobility patterns of HCWs and motivational factors, calling for systematic data and research evidence.
- The major push and pull factors reveal strong interconnectedness of healthcare systems and policy in sending and receiving countries, calling for transnational EU solutions and solidarity-based global HCWF policy.
- The capacity for circular migration must be explored systematically. Connecting bottom-up motivation and institutional conditions may reveal new pathways of effective and equitable HCWF policy.

## Data Availability

The raw data supporting the conclusions of this article are not publicly available to ensure anonymity of the interview participants, but will be made available by the authors on reasonable request.

## Acknowledegments

We thank the participants in the interview study for sharing their experiences and making time for supporting our research even in a period of a very high workload. We also thank Frank Müller, the leading project partner at the University Medical Centre Göttingen.

## Ethics statement

The qualitative interview study was approved by the Ethics Committee of Hannover Medical School (8973_BO_K_2020; 9948_BO_K_2021) and University Medical Center Göttingen (Nr. 39/8/21). Informed consent was obtained from all physicians participating in the interviews.

## Author contributions

EK and AD-J had the idea and supervised the research; EK, AJ-D and M-IU developed the design; M-IU, NCC, MGB provided expert advice on Romania; EK gathered the interviews; NT and LMF supported data collection and analysis; EK, M-IU and MGB organised webinars and an international conference session; EK prepared a first draft; all authors provided comments and have read and approved the final manuscript.

